# Tracking the Changes in Longitudinal MRI-detected Perivascular Spaces following Ischaemic Stroke

**DOI:** 10.64898/2026.03.16.26348475

**Authors:** William Pham, Mohamed Salah Khlif, Zhibin Chen, Alexander Jarema, Luke A. Henderson, Vaughan G. Macefield, Amy Brodtmann

## Abstract

Stroke is a leading cause of mortality and morbidity worldwide. MRI-visible perivascular spaces (PVS) are an emerging marker of cerebral small vessel disease and may have prognostic value in stroke. We investigated longitudinal changes in PVS volume and cluster count following ischaemic stroke. PVS volumes and cluster counts were compared between stroke survivors (*n*=124; 39 women; median [Q1, Q3] age=70 [62, 76] years) and healthy controls (*n*=39; 15 women; median age=69 [66, 72.5] years). MRI scans were acquired at 3 months, 12 months, and 36 months post-stroke. PVS were automatically segmented from T1-weighted MRI using a validated deep learning algorithm (nnU-Net). Generalised linear mixed-effects models were used to assess group differences and longitudinal changes in PVS, adjusting for baseline age, sex, total intracranial volume, and BMI. At the 12-month timepoint, no significant differences in PVS metrics were observed between stroke and control groups. However, at the 36-month timepoint we observed a significant brain-wide reduction in PVS volume (exp(β)=0.93, 95%CI [0.87, 1], *p*=0.035) and cluster count (exp(β)=0.92, 95%CI [0.85, 0.99], *p*=0.003) in the stroke group compared to control. Regionally, by 36 months, stroke patients demonstrated significant PVS reductions relative to controls in the frontal (PVS volume: exp(β)=0.93, 95%CI [0.82, 0.99], *p*=0.032; PVS cluster counts: exp(β)=0.91, 95%CI [0.83, 1], *p*=0.037) and parietal lobes (PVS volume: exp(β)=0.93, 95%CI [0.85, 1.01], *p*=0.10; PVS cluster counts: exp(β)=0.84, 95%CI [0.68, 1.08], *p*<0.001). These findings suggest that ischaemic stroke is associated with dynamic and regional changes in PVS volume and counts.

## Introduction

Ischaemic stroke accounts for 71% of all stroke cases and is a leading cause of worldwide mortality and morbidity (Brauer et al., 2024; Campbell et al., 2019). Ischaemic stroke is associated with accelerated brain-wide atrophy, cortical thinning, and white matter neurodegeneration (Brodtmann et al., 2012, 2020; Brodtmann, Khlif, et al., 2021; Egorova-Brumley et al., 2023).These structural changes evolve over months to years following stroke.

Perivascular spaces (PVS) are fluid-filled spaces surrounding cerebral blood vessels. According to the Standards for Neuroimaging Research in Small Vessel Disease (STRIVE) criteria, enlarged PVS are identified as key MRI markers of cerebral small vessels disease (CSVD) along with lacunar infarctions, white matter hyperintensities (WMH), cerebral microbleeds, and cortical superficial siderosis (Duering et al., 2023). Individuals with CSVD are at elevated risk of both ischaemic stroke and subsequent cognitive decline (Lau, Li, Schulz, et al., 2017). MRI-visible PVS are increasingly regarded as non-invasive biomarkers of CSVD, with enlargement linked to ageing and cerebrovascular pathology (Duperron et al., 2018; Lynch et al., 2023).

Perivascular spaces also function as critical pathways for cerebrospinal fluid movement and waste clearance within the glymphatic system (Iliff et al., 2012). Animal studies have shown that the perivascular fluid flow along the glymphatic system is a major driver of cerebral oedema post-ischaemia (Mestre et al., 2020). Moreover, impaired glymphatic function after ischaemic stroke has been linked to the deposition of amyloid-exp(β) oligomers and secondary neuronal injury in the ipsilateral thalamus (Gu et al., 2025).

Traditionally, investigations of perivascular space enlargement in MRI relied on visual rating, where enlargement scores are assigned based on the number of visible PVS clusters in selected axial slices and on predefined thresholds (Potter et al., 2015). Accordingly, PVS enlargement in the centrum semiovale has been identified as a predictor of early mortality in haemorrhagic stroke (Song et al., 2021). Whereas, basal ganglia (BG) PVS counts are associated with an increased risk of haemorrhagic rebleeding (Tian et al., 2024). No significant associations between PVS burden in the BG or centrum semiovale and the risk of ischaemic stroke recurrence were found in one meta-analysis (Lei et al., 2024). This is possibly due visual rating of PVS being suboptimal, reliant on predefined slices and subjective assessments, and therefore fails to capture the full extent of PVS enlargement.

Recent advances in MRI segmentation algorithms have enabled voxel-wise quantification of PVS throughout the entire brain (Pham et al., 2022; Waymont et al., 2024). In this study, we utilised our own deep learning models to automatically quantify PVS (volume and cluster count) across several brain regions based on T1-weighted MRI scans. We aimed to track the longitudinal PVS changes in ischaemic stroke survivors scanned 3-, 12-, and 36-months post-stroke. We hypothesised that stroke patients would exhibit greater PVS burden than stroke-free controls, with greater PVS enlargement (i.e., increased counts and volume) over time.

## Methods

The *Cognition and Neocortical Volume After Stroke* (CANVAS) study is a prospective observational cohort study comprising individuals with ischaemic stroke in any site or territory (*n*=124) and age– and sex-matched controls without a history of stroke (*n=*39) (Brodtmann et al., 2014). Participants were recruited from three hospitals in Victoria, Australia: Austin Health, Box Hill Hospital, and the Royal Melbourne Hospital. Stroke patients were enrolled from the Acute Stroke Units at each site, with diagnosis confirmed through clinical neuroimaging (CT or MRI). Exclusion criteria for both groups included a history of cognitive impairment, inability to undergo MRI scanning, or a diagnosis of a severe medical illness. Assessments were conducted at up to 5 timepoints: baseline<6 weeks of stroke, 3-, 12-, 36– and 60-months post-stroke.

The study was approved by respective Human Research Ethics Committees in participating institutions. Informed written consent was obtained from all participants in accordance with the Declaration of Helsinki.

## Participants

We included participants with available data at 3-, 12– and 36-months for this analysis. Demographic and clinical information were collected including age, sex, body mass index (BMI), vascular risk factors (VRFs), and comorbidities.

## Imaging protocol

Whole-brain imaging was performed using a 3T Siemens Tim Trio scanner equipped with a 12-channel head coil (Siemens, Erlangen, Germany). T1-weighted (T1w) images were acquired using a magnetisation-prepared rapid acquisition gradient echo (MPRAGE) sequence with the following parameters: TR=1900 ms; TE=2.55 ms; TI=900 ms; FA=9°; FOV=256 × 256; and voxel size=1.0 mm^3^ isotropic. All scans were manually reviewed and images affected by motion artefacts or other sources of degradation were excluded.

## Perivascular space quantification

PVS were automatically segmented in T1w MRI scans using a previously validated convolutional neural network (nnU-Net) (Isensee et al., 2021; Pham et al., 2025). The model produced segmentation maps with distinct voxel-wise labels for PVS in the white matter and basal ganglia. Midbrain and hippocampal PVS were automatically quantified by specialised nnU-Net models (Pham et al., 2025). The two PVS metrics considered in this study were volumes and cluster counts. PVS enlargement was defined as an increase in either PVS volume or count, and PVS shrinkage as a decrease in either metric. PVS cluster counts were determined using the ‘measure.label’ function from the ‘scikit-image’ Python package (Walt et al., 2014), which identifies spatially distinct PVS clusters within the segmentation mask. All PVS segmentation, processing and quantification were conducted using Python (v3.9.13).

### Quantification via region and arterial territory

PVS metrics were further quantified within specific regions of interest (ROIs). A total of seven ROIs were defined based on two brain atlases. Lobar regions, including the frontal, parietal, temporal, and occipital lobes, were defined according to the ICBM2009 asymmetrical atlas (Fonov et al., 2009). Additionally, PVS metrics were quantified within supratentorial cerebral arterial territories, including the anterior cerebral artery (ACA), middle cerebral artery (MCA), and posterior cerebral artery (PCA) regions, as defined by a vascular territory atlas (Liu et al., 2023).

While the lobar regions reflect the functional organisation at a whole-brain level, vascular territories delineate areas with distinct blood perfusion supplied by major arterial branches. Substantial overlap exists between these two parcellations (Figure 1). The ACA supplies the medial portions of the frontal and parietal lobes, the MCA perfuses the lateral aspects of all four lobes, and the PCA primarily supplies the occipital lobe and posterior temporal lobe.

**Figure 1.**
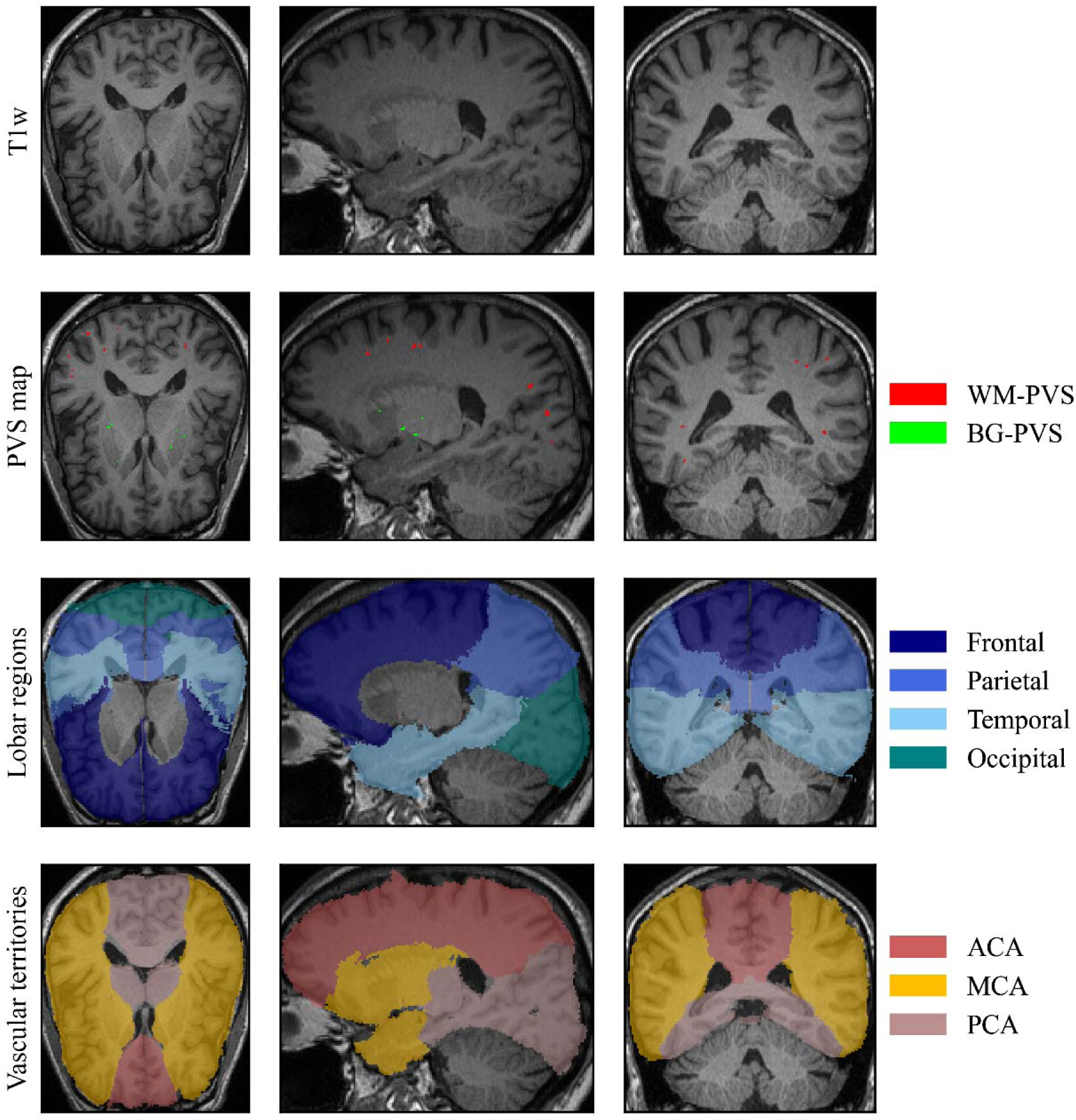
Examples of T1-weighted (T1w) MRI with segmented perivascular spaces (PVS) maps and region atlas parcellations according to lobar regions and arterial vascular territories. Top row: Raw T1w MRI scan from a stroke-free participant with images from the axial, sagittal, and coronal planes. Second row: PVS map overlaid on T1w MRI, with white matter PVS (WM-PVS, red) and basal ganglia PVS (BG-PVS, green). Third row: Lobar regions including the frontal (dark blue), parietal (light blue), temporal (cyan), and occipital (teal) lobes. Bottom row: Arterial vascular territories including the regions supplied by the anterior cerebral artery (ACA, brown), middle cerebral artery (MCA, yellow), and posterior cerebral artery (PCA, pink).

T1w MRI scans were further processed using FastSurfer (v2) to extract baseline total intracranial volume (TIV) and generate brain parcellations for each subject (Desikan et al., 2006; Henschel et al., 2020). Extracted brain volumes were spatially registered to the MNI152 template using Advanced Normalization Tools (ANTs, v2.4.3) (Avants et al., 2011). Registration was performed with antsRegistrationSyn.sh (rigid, affine, symmetric diffeomorphic normalisation [SyN]), producing forward and inverse transforms. Subject-specific atlases were generated by applying the inverse warp fields and affine transformations to the ICBM2009c atlas and arterial territories atlas.

PVS volumes and cluster counts were computed for each ROI by overlaying the subject-specific atlases with corresponding PVS segmentation maps. A PVS cluster was considered to belong to a given ROI if its centroid was situated within that ROI. The centroid was determined with the sci-kit image ‘measure.centroid’ function. The nnU-Net model was trained to specifically segment perivascular spaces and not stroke lesions.

All image processing was performed on the MASSIVE High Performance Computing cluster (Goscinski et al., 2014). Perivascular space segmentation using nnU-Net was conducted on an NVIDIA A40 GPU equipped with 256 GB RAM.

## Statistical Analysis

To assess the reliability of the automated PVS measurements, a validation procedure was conducted using manual PVS counts as the reference standard. In the baseline T1w MRI of all subjects, PVS were manually counted in two representative axial slices in the centrum semiovale (CS) and basal ganglia (BG). In the BG, one axial slice was positioned at the level of the anterior commissure and the second axial slice superior to the anterior commissure. PVS were also manually counted throughout the entire midbrain and hippocampal regions. For the CS and BG, automated PVS counts were extracted from the same axial slices used for manual rating, while full-region comparisons were performed for the midbrain and hippocampi.

We computed Lin’s concordance correlation coefficient (CCC) to evaluate the agreement between manual and automated PVS counts, and Spearman’s rank correlation coefficient to assess the monotonic association between the two methods. PVS validation was conducted on all baseline MRI scans. Manual annotations were performed by an experienced rater (WP) and independently reviewed by a board-certified radiologist (AJ).

We assessed group differences in PVS metrics and their longitudinal changes using generalised linear mixed-effects models (GLMMs). Since most PVS metrics were not normally distributed, we modelled them using a Tweedie distribution with a log link function. GLMMs with the Tweedie distribution demonstrated good model fit for all PVS metrics, except for those in the midbrain and hippocampus. Consequently, PVS metrics in the midbrain and hippocampus were binarised into ‘high’ and ‘low’ categories based on a median cut-off. Binary outcomes were modelled using GLMMs with binomial family and log-link functions to evaluate the effects of stroke diagnosis and the group × timepoint interaction term.

All models included PVS metrics as the dependent variable, with timepoint × group interaction as a fixed effect, and a random intercept for each subject to account for intra-individual differences. Baseline age, sex, TIV, and BMI were included as covariates in all models as these factors have previously been associated with PVS measurements (Barisano et al., 2021; Francis et al., 2019). Since previous studies have shown age-related changes in PVS to be non-linear, baseline age was modelled using a second-order spline term (Lynch et al., 2023; Park et al., 2023).

For all GLMMs, the normality of residuals was assessed by visually inspecting Q-Q plots, and homoscedasticity was assessed by inspecting the residual plots. Model diagnostics included visual inspection of residuals, assessment of overdispersion and zero-inflation using the ‘simulateResiduals’ function from the *DHARMa* package, and comparison of Akaike Information Criterion (AIC) and Bayesian Information Criterion (BIC) for model selection. Covariates were removed if inclusion compromised residual normality or homoscedasticity.

To derive robust estimates, 95% confidence intervals (CIs) for fixed effects were calculated using parametric bootstrapping with 1,000 simulations. Statistical significance was assessed with an α level of 0.05. All GLMMs were implemented using the *glmmTMB* package in R (v4.2.1). To account for multiple comparisons, *p*-values from all models were adjusted using the Benjamini-Hochberg false discovery rate (FDR) correction.

## Results

One hundred and sixty-three participants (54 females, median [Q1, Q3] age=69 [63, 75] years) had available data: 124 stroke survivors (stroke group) and 39 stroke-free controls (control group). Groups were balanced for age and sex, but the control had significantly more years of education and, unsurprisingly, the stroke group had more vascular risk factors. Across the cohort, median [Q1, Q3] total PVS volume and cluster counts at baseline were 1,655 [1,094, 2,263] mm^3^ and 153 [108, 205.5], respectively. There was no significant difference in baseline PVS volume and cluster counts between groups. Subject demographics and clinical characteristics are summarised in Table 1.

**Table 1.** Baseline demographic and clinical characteristics of the study cohort. For normally distributed numerical variables, values are presented as mean ± SD. For non-normally distributed numerical variables, values are presented as median [Q1, Q3]. TIV=total intracranial volume; VRF=vascular risk factors; PVS=perivascular space; BMI=body mass index. Categorical variables were compared using the Chi-squared test, and continuous variables using independent samples t-tests.

| Characteristic | Control (n=39) | Stroke (n=124) | Total (N=163) | p-value |
| --- | --- | --- | --- | --- |
| Age (years) | 69 [66, 72.5] | 70 [62, 76] | 69 [63, 75] | 0.79 |
| Sex, n female (%) | 15 (38.5%) | 39 (31.5%) | 54 (33.1%) | 0.42 |
| Education (years) | 17 [11, 18] | 12 [10, 15] | 13 [10, 16] | <0.001 |
| Baseline TIV (cm<sup>3</sup>) | 1,471.8 ± 112.9 | 1,449.3 ± 155.6 | 1,454.7 ± 146.5 | 0.41 |
| BMI | 26.6 ± 3.8 | 27.7 ± 4.4 | 27.4 ± 4.2 | 0.14 |
| Composite VRFs, n (%) | 16 (41%) | 87 (70.2%) | 103 (63.2%) | <0.001 |
| PVS Volume (mm<sup>3</sup>) | 1,469 [989, 2,020] | 1,727.5 [1,134.3, 2,493.3] | 1,655 [1,094, 2,263] | 0.09 |
| PVS Clusters | 144 [93.5, 178.5] | 159.5 [109.5, 218.5] | 153 [108, 205.5] | 0.10 |

## Regional validation of automated perivascular space quantification

Agreement between manual and automated PVS counts was evaluated in selected axial slices for the CS and BG, as well as across the entire midbrain and hippocampal regions. Strong agreement was observed in the CS, with a high correlation and concordance between manual and predicted counts (Spearman’s ρ=0.84, *p* < 0.001; CCC=0.76, 95%CI [0.73, 0.79]). Low agreement was found for the BG-PVS counts (Spearman’s ρ=0.67, *p* < 0.001; CCC=0.57, 95%CI [0.53, 0.61]). The midbrain PVS counts showed weak correlation and low concordance between manual and predicted counts (Spearman’s ρ=0.54, *p* < 0.001; CCC=0.53, 95%CI [0.41, 0.63]), while the hippocampal PVS counts demonstrated moderate correlation and agreement (Spearman’s ρ=0.72, *p* < 0.001; CCC=0.68, 95%CI [0.6, 0.74]). Notably these patterns of agreement were consistent across both stroke patients and controls.

## Global perivascular space comparison

At baseline, the median total PVS volume was higher in stroke patients (1,723 [1,134.3, 2,493.3] mm^3^) than in controls (1,451 [989, 2,020] mm^3^). After adjusting for covariates, stroke group exhibited significantly greater total PVS volume (exp(β)=1.38, 95% CI [1.13, 1.69], *p*=0.003, *p*_FDR_=0.015) and total PVS cluster count (exp(β)=1.34, 95% CI [1.09, 1.63], *p*=0.003, *p*_FDR_=0.015). While controls exhibited progressive enlargement of whole-brain PVS volume and cluster counts across the 3 years, stroke patients demonstrated significant shrinkage over the same period. At 36 months, stroke group showed significantly greater reductions in both PVS volume (exp(β)=0.93, 95% CI [0.87, 1], *p*=0.035, *p*_FDR_=0.092) and cluster count (exp(β)=0.92, 95% CI [0.85, 0.99], *p*=0.02, *p*_FDR_=0.063) compared to control group. No significant between-group effects were observed at the 12-month timepoint for any global or regional PVS metric (all *p* > 0.05). All model results are summarised in **Error! Reference source not found.** and illustrated in **Error! Reference source not found.**-2.

## Perivascular spaces in the white matter and basal ganglia

In the white matter, both PVS volume (exp(β)=1.58, 95% CI [1.19, 2.1], *p*=0.001, *p*_FDR_=0.009) and cluster count (exp(β)=1.41, 95% CI [1.1, 1.82], *p*=0.004, *p*_FDR_=0.018) were significantly higher in stroke group. Over 36 months, the stroke group exhibited significantly greater reductions in WM PVS volume (exp(β)=0.91, 95% CI [0.84, 0.99], *p*=0.037, *p*_FDR_=0.092) and cluster count (exp(β)=0.91, 95% CI [0.83, 0.98], *p*=0.021, *p*_FDR_=0.063) in the stroke group across the 36-month period compared to controls.

In contrast, there was no significant group effect for BG PVS volume (exp(β)=1.11 95% CI [0.94, 1.29], *p*=0.21), nor significant group by time interaction at the 36-month timepoint (exp(β)=0.96, 95% CI [0.9, 1.02], *p*=0.22). BG PVS cluster counts were significantly greater in the stroke group compared to controls (exp(β)=1.19, 95% CI [1.04, 1.39], *p*=0.019, *p*_FDR_=0.062), however, there were no significant between-group effects at the 36-month timepoint for BG PVS cluster counts (exp(β)=0.93, 95% CI [0.86, 1.02], *p*=0.11).

## Lobar perivascular spaces

The stroke group exhibited significantly higher PVS volume and cluster counts in the frontal (volume: exp(β)=1.69, 95% CI [1.28, 2.33], *p*=0.001, *p*_FDR_=0.009; counts: exp(β)=1.43, 95% CI [1.12, 1.84], *p*=0.003, *p*_FDR_=0.015), parietal (volume: exp(β)=1.39, 95% CI [1.04, 1.83], *p*=0.019, *p*_FDR_=0.062; counts: exp(β)=1.69, 95% CI [0.78, 4.95], *p*<0.001, *p*_FDR_<0.001), temporal (volume: exp(β)=1.6, 95% CI [1.12, 2.31], *p*=0.008, *p*_FDR_=0.03; counts: exp(β)=1.52, 95% CI [1.12, 2.04], *p*=0.005, *p*_FDR_=0.021) than controls. In the occipital lobe, neither PVS volumes (exp(β)=1.61, 95% CI [0.82, 3.12], *p*=0.18) nor PVS counts (exp(β)=1.26, 95% CI [0.08, 21.33], *p*=0.84) were significantly different between stroke and controls.

During the 3-year period, parietal lobe PVS volume and counts increased in controls. By contrast, the stroke group showed significant reductions in parietal lobe PVS cluster counts (exp(β)=0.84, 95% CI [0.68, 1.08], *p*<0.001, *p*_FDR_<0.001) but not PVS volumes (exp(β)=0.93, 95% CI [0.85, 1.01], *p*=0.10). No significant group by time interactions were observed in the temporal lobe for either PVS volume or cluster counts (all *p* > 0.05).

In the frontal lobe, both PVS volumes (exp(β)=0.9, 95% CI [0.82, 0.99], *p*=0.032, *p*_FDR_=0.089) and cluster counts (exp(β)=0.91, 95% CI [0.83, 1], *p*=0.037, *p*_FDR_=0.092) decreased significantly in the stroke patients relative to controls over the 36-month period. In contrast, PVS metrics in the temporal (volume: exp(β)=0.95, 95% CI [0.86, 1.01], *p*=0.10; counts: exp(β)=0.93, 95% CI [0.83, 1.03], *p*=0.17) and occipital lobes (volume: exp(β)=0.9, 95% CI [0.78, 1.06], *p*=0.23; counts: exp(β)=1.11, 95% CI [0.18, 4.96], *p*=0.44) showed no significant difference to controls over the 36-month period.

## Perivascular spaces via vascular territories

The vascular territory-based analyses largely reflect the same underlying findings as the lobar analyses. This overlap arises because the MCA perfuses most cortical and subcortical being examined in our study. Nevertheless, we observed that PVS volume was significantly higher at baseline in the stroke group than the control group in the ACA (exp(β)=1.59, 95% CI [1.17, 2.12], *p*=0.003, *p*_FDR_=0.015) and MCA territories (exp(β)=1.32, 95% CI [1.1, 1.62], *p*=0.004, *p*_FDR_=0.018), but not PCA territory (exp(β)=1.28, 95% CI [0.88, 1.85], *p*=0.19). Similarly, PVS cluster counts were significantly higher in the stroke group than control in the ACA (exp(β)=1.36, 95% CI [1.07, 1.75], *p*=0.014, *p*_FDR_=0.05), MCA (exp(β)=1.34, 95% CI [1.1, 1.59], *p*=0.002, *p*_FDR_=0.014), and PCA territories (exp(β)=6.39, 95% CI [0.04, 2036.52], *p*<0.001, *p*_FDR_<0.001).

Over the 36-month period, stroke-free controls exhibited PVS enlargement whereas stroke patients exhibited PVS shrinkage in the MCA territory (volume: exp(β)=0.93, 95% CI [0.87, 0.99], *p*=0.023, *p*_FDR_=0.066; counts: exp(β)=0.9, 95% CI [0.84, 0.97], *p*=0.006, *p*_FDR_=0.024). No significant longitudinal differences in PVS metrics in the ACA vascular territory were observed between groups over the 36-month period (all *p* > 0.05).

## Perivascular spaces in the midbrain and hippocampus

The hippocampus is largely supplied by the PCA, whereas the midbrain is located within the posterior regions but is not supplied by the PCA. The logistic mixed-effects models revealed no significant group effects or longitudinal changes in midbrain or hippocampal PVS volumes or cluster counts (all *p* > 0.05).

**Figure 2.**
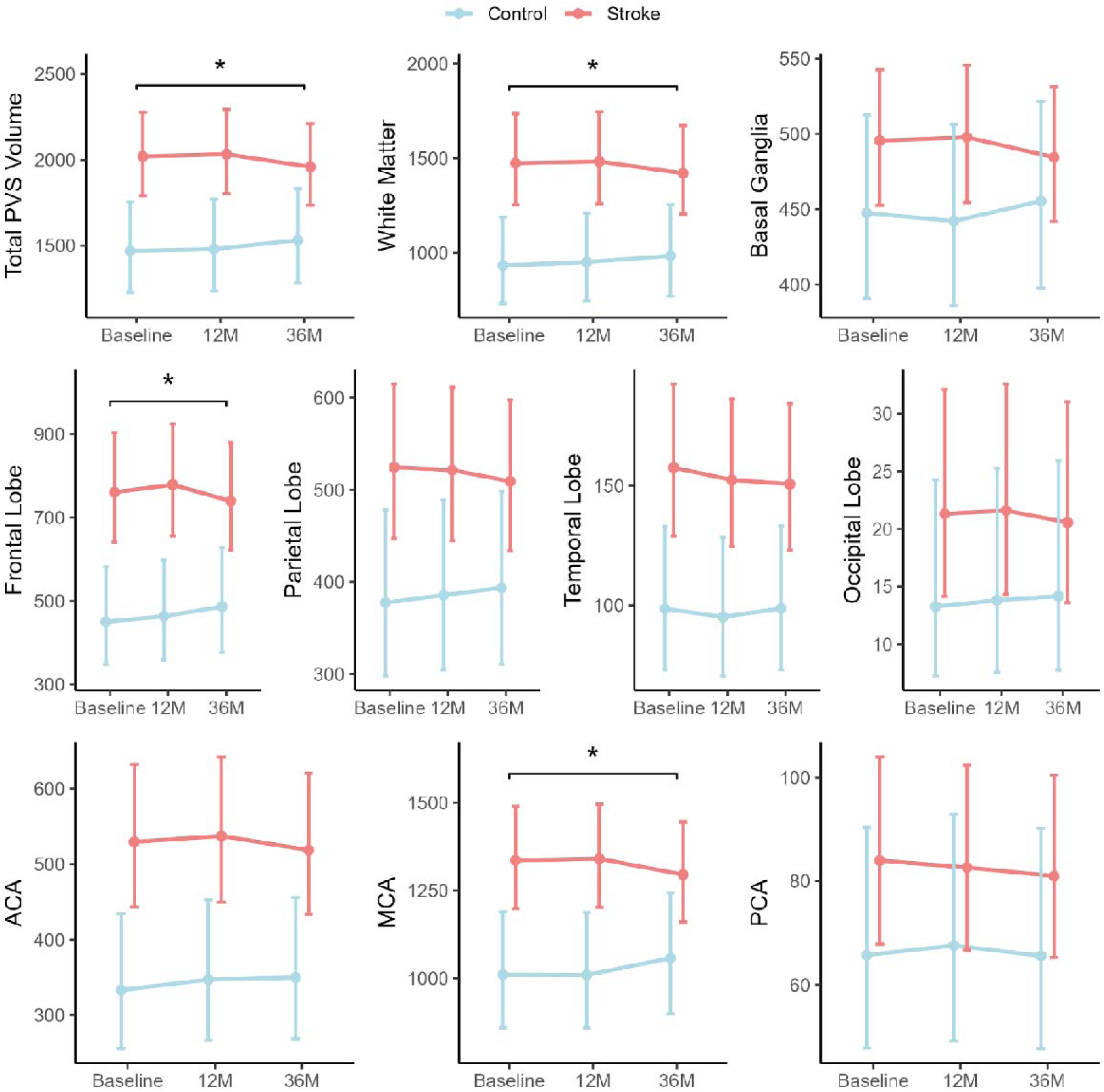
Longitudinal changes in predicted perivascular space volumes by brain region and arterial territory in stroke patients versus controls. Line plots display estimated marginal means (±95% CI) from generalised linear mixed effects models for total, regional, and vascular territory PVS volumes across three timepoints: baseline, 12 months, and 36 months. Blue lines represent controls; red lines represent stroke patients. Significant group effects and Group × Time (36M) interaction effects are annotated in each panel with corresponding exp(β) estimates and bootstrapped 95% CIs. Significance: *p<0.05, **p<0.01, ***p<0.001. Significant PVS volume reductions were observed in stroke patients at the 36-month timepoint across the whole brain and in several regions including the white matter, frontal lobe, and middle cerebral artery territory. However, none of these associations remained

**Figure 3.**
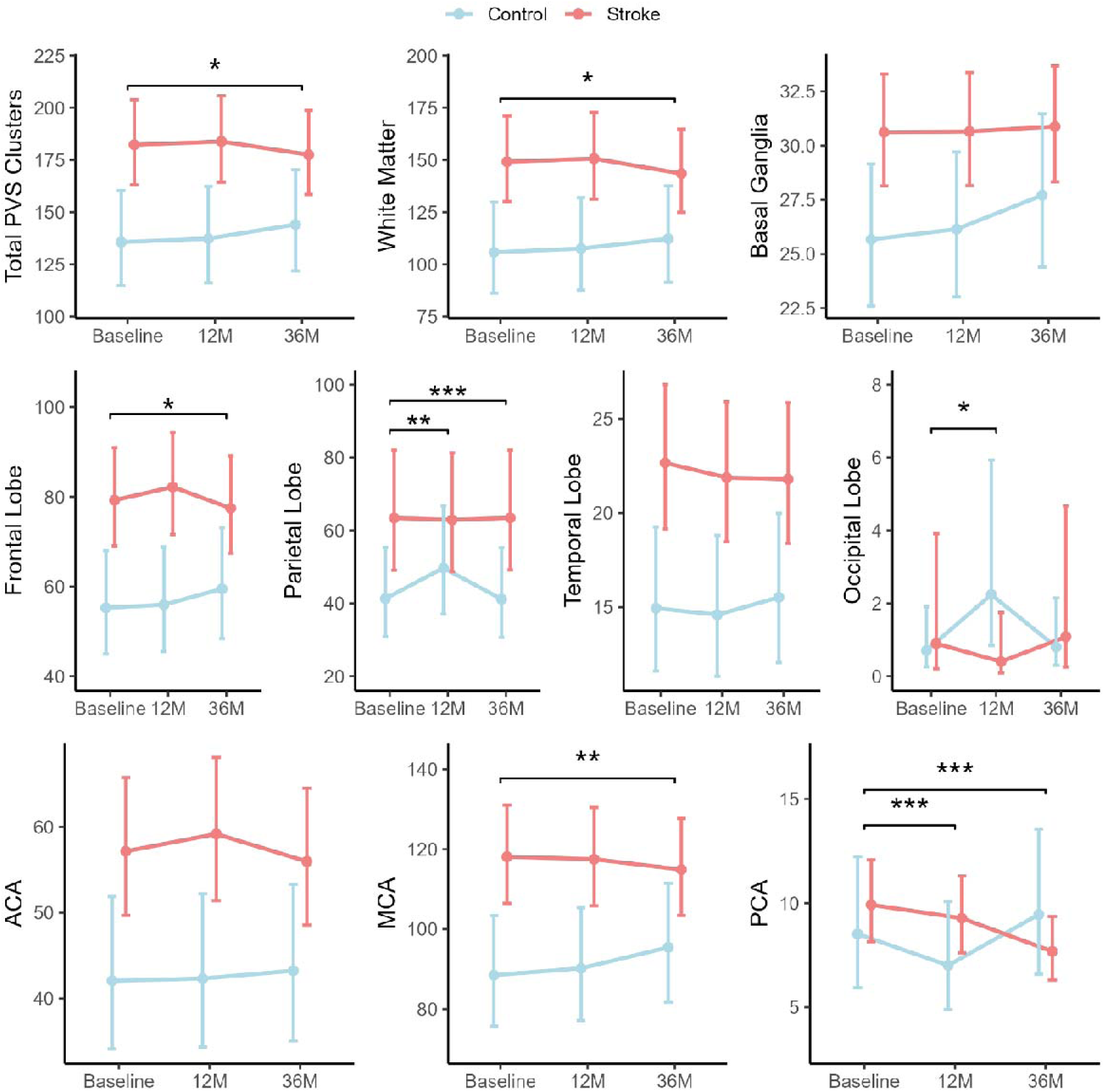
Longitudinal changes in predicted perivascular space cluster counts by brain region and arterial territory in stroke versus control groups. Line plots display estimated marginal means (±95% CI) from generalised linear mixed effects models for total, regional, and vascular territory PVS cluster counts across three timepoints: baseline, 12 months, and 36 months. Blue lines represent controls; red lines represent stroke patients. Significant group effects and Group × Time (36M) interaction effects are annotated in each panel with corresponding exp(β) estimates and bootstrapped 95% CIs. Significance: *p<0.05, **p<0.01, ***p<0.001. Significant reductions in PVS cluster counts were observed in stroke patients at the 36-month timepoint across the whole brain and in several regions including the white matter, frontal and parietal lobes, and middle and posterior cerebral artery territories. After correcting for multiple comparisons, associations in the parietal lobe and the middle and posterior cerebral artery territories remained statistically significant (*p*_FDR_<0.05). ACA: anterior cerebral artery. MCA: middle cerebral artery. PCA: posterior cerebral artery.

## Discussion

In this longitudinal case-control study, we observed spatially distinct alterations in MRI-visible perivascular spaces in the ischaemic stroke group. The stroke group exhibited significantly enlarged PVS compared to the control group in the white matter, frontal, parietal and temporal lobes, and anterior and middle cerebral artery territories. Over the study period, PVS counts and volumes increased in the control group, consistent with age-related enlargement. Contrary to our hypothesis, stroke group PVS volume and counts reduced over the 36-month period, with the most pronounced reductions observed in the parietal lobe and MCA territory, compared to baseline. Together, the findings indicate that ischaemic stroke induces region-specific changes in perivascular spaces, with stroke patients showing a reduction in perivascular spaces within the first three years post-stroke.

Previous studies investigating PVS in stroke populations have yielded inconsistent results, often using semi-quantitative 2D rating scales (Ramaswamy et al., 2022). Enlarged basal ganglia PVS have been identified in two studies as a risk factor for incident ischaemic stroke (Lau, Li, Lovelock, et al., 2017; Lau, Li, Schulz, et al., 2017), but an association was found between BG PVS and lacunar ischaemic stroke only in another study (Doubal et al., 2010). A meta-analysis reported significant associations between PVS burden and ageing, hypertension, and lacunes, but found no direct association with stroke (Francis et al., 2019). In contrast, Ballerini et al., (2020) applied computational methods to quantify perivascular spaces in community-dwelling individuals and observed significantly increased PVS width and size in participants with recurrent stroke (Ballerini et al., 2020).

Our finding of a longitudinal reduction in PVS count and volume in stroke survivors is novel. This observation was made possible by our longitudinal study design, which followed patients over a three-year period, and has implications for the interpretation of MRI-perivascular space measurements. Currently, the MRI-PVS enlargement is regarded an biomarker of CSVD (Duering et al., 2023). Consistent with this idea, our stroke group exhibited a significantly larger PVS burden than controls; however, the longitudinal PVS trajectories progressed in opposite directions between the two groups: over the three-year study period, perivascular spaces continued to enlarge in controls, whereas stroke patients demonstrated significant shrinkage in multiple brain regions.

Enlarged PVS in stroke patients may reflect pathological processes preceding the ischaemic event or may instead result from the stroke itself and associated hyperacute patho-physiological mechanisms. For example, hyperacute enlargement of perivascular spaces can be driven by mechanisms such as impaired glymphatic clearance, blood-brain-barrier dysfunction, and inflammatory cell infiltration (Hosoki et al., 2023; Mestre et al., 2020). Our study design does not allow us to determine whether PVS changes precede stroke, either as a causal factor or epiphenomenon, or whether they arise as a consequence of the stroke event.

Nevertheless, our finding of PVS shrinkage three years after ischaemic stroke supports the notion that PVS volume and count trajectories are not linear (Lynch et al., 2023), and that PVS morphology alone may not serve as a readily interpretable biomarker of brain health. It is possible that accelerated brain atrophy, which we previously reported in this cohort (Brodtmann et al., 2020; Brodtmann, Werden, et al., 2021) would have contributed to structural collapse of white matter, thereby reducing the volume and visibility of PVS.

Furthermore, we found no significant associations between stroke diagnosis and PVS burden in the midbrain or hippocampus. The hippocampus is a largely PCA-supplied region, and the midbrain is supplied by posterior circulation (basilar) branches below the circle of Willis arteries examined here (Johnson, 2023). Enlarged PVS in the midbrain has been linked to Parkinson’s disease and cerebrospinal fluid levels of tau protein (Li et al., 2020). In a previous study, we observed significant ipsilesional atrophy of hippocampal subfields in ischaemic stroke patients when assessed at the 12-month follow-up (Khlif et al., 2022). Given the small sample size and lower prevalence of detectable PVS in midbrain and hippocampi, larger cohorts may be necessary to detect subtle PVS changes in these regions.

## Strengths and limitations

Strengths of this study include the use of a longitudinal design and a novel validated deep learning approach for automated PVS quantification. However, several limitations should be acknowledged. First, our cohort was restricted to patients with ischaemic stroke, limiting generalisability to other stroke subtypes such as haemorrhagic stroke. Second, stroke heterogeneity, including variation in lesion size, location, arterial territory, aetiology, and side could have introduced variability. We did not investigate whether the stroke side or site influenced the spatiotemporal evolution of MRI-visible PVS.

Further, we did not examine associations between PVS burden and other markers of CSVD, such as WMH, lacunar infarctions or cerebral microbleeds. The presence of WMH can obscure the visibility of PVS in MRI (Wardlaw et al., 2013), and previous studies have reported that microbleeds often co-occur with enlarged PVS (Bouvy et al., 2016, 2020). Finally, PVS were segmented from T1-weighted MRI, which is inherently less sensitive for PVS detection compared to T2-weighted sequences.

## Conclusion

We found that stroke survivors exhibited greater PVS counts and volumes compared to stroke-free controls at the 3-month timepoint following stroke. However, while the control group exhibited an expected age-related increase in PVS count and volume over the three-year follow-up period, our stroke group exhibited reductions in PVS count and volumes in several brain regions. This resulted in significant between-group differences in PVS count and volume over time. PVS count and volume may capture cerebrovascular changes associated with chronic brain remodelling. In summary, we provide novel evidence that perivascular space alterations are dynamic over the 3 years after ischaemic stroke. While these observations highlight the potential of MRI-PVS as prognostic imaging biomarkers of post-stroke outcomes, further research is necessary to clarify their biological underpinnings and determine the clinical contexts in which PVS measurements could be most effectively applied.

## Data Availability

The dataset analysed in the current study are available from the corresponding author upon reasonable request.

## Acknowledgements

The authors thank the University of Melbourne Victorian Life Sciences Computation Initiative, Florey Node National Imaging Facility, Melbourne Brain Centre radiographers, and all our participants, who so generously contributed their time to the study.

## Funding

Financial support for this study was received from: NHMRC GNT1020526, GNT1045617 (AB), GNT1094974; Brain Foundation; Wicking Trust; Collie Trust; Sidney and Fiona Myer Family Foundation; Australian Research Council DE180100893 (NE); and Heart Foundation Future Leader Fellowship 100784 (AB).

## Notes

**Support:** This research was supported by an Australian Government Research Training Program (RTP) Scholarship.

### Competing Interest Statement

The authors have declared no competing interest.

### Author Declarations

Participants were recruited from three hospitals in Victoria, Australia: Austin Health, Box Hill Hospital, and the Royal Melbourne Hospital. The study was approved by respective Human Research Ethics Committees in participating institutions. Informed written consent was obtained from all participants in accordance with the Declaration of Helsinki.

## References

1. Avants, B. B., Tustison, N. J., Song, G., Cook, P. A., Klein, A., & Gee, J. C. (2011). A reproducible evaluation of ANTs similarity metric performance in brain image registration. NeuroImage, 54(3), 2033–2044. 10.1016/j.neuroimage.2010.09.025

2. Ballerini, L., Booth, T., Valdés Hernández, M. del C., Wiseman, S., Lovreglio, R., Muñoz Maniega, S., Morris, Z., Pattie, A., Corley, J., Gow, A., Bastin, M. E., Deary, I. J., & Wardlaw, J. (2020). Computational quantification of brain perivascular space morphologies: Associations with vascular risk factors and white matter hyperintensities. A study in the Lothian Birth Cohort 1936. NeuroImage: Clinical, 25. 10.1016/j.nicl.2019.102120

3. Barisano, G., Sheikh-Bahaei, N., Law, M., Toga, A. W., & Sepehrband, F. (2021). Body mass index, time of day and genetics affect perivascular spaces in the white matter. Journal of Cerebral Blood Flow & Metabolism, 41(7), 1563–1578. 10.1177/0271678X20972856

4. Bouvy, W. H., van Veluw, S. J., Kuijf, H. J., Zwanenburg, J. J. M., Kappelle, J. L., Luijten, P. R., Koek, H. L., Geerlings, M. I., & Biessels, G. J. (2020). Microbleeds colocalize with enlarged juxtacortical perivascular spaces in amnestic mild cognitive impairment and early Alzheimer’s disease: A 7 Tesla MRI study. Journal of Cerebral Blood Flow and Metabolism, 40(4), 739–746. 10.1177/0271678X19838087

5. Bouvy, W. H., Zwanenburg, J. J. M., Reinink, R., Wisse, L. E. M., Luijten, P. R., Kappelle, L. J., Geerlings, M. I., & Biessels, G. J. (2016). Perivascular spaces on 7 Tesla brain MRI are related to markers of small vessel disease but not to age or cardiovascular risk factors. Journal of Cerebral Blood Flow and Metabolism, 36(10), 1708–1717. 10.1177/0271678X16648970

6. Brauer, M., Roth, G. A., Aravkin, A. Y., Zheng, P., Abate, K. H., Abate, Y. H., Abbafati, C., Abbasgholizadeh, R., Abbasi, M. A., Abbasian, M., Abbasifard, M., Abbasi-Kangevari, M., Abd ElHafeez, S., Abd-Elsalam, S., Abdi, P., Abdollahi, M., Abdoun, M., Abdulah, D. M., Abdullahi, A., … Gakidou, E. (2024). Global burden and strength of evidence for 88 risk factors in 204 countries and 811 subnational locations, 1990–2021: A systematic analysis for the Global Burden of Disease Study 2021. The Lancet, 403(10440), 2162–2203. 10.1016/S0140-6736(24)00933-4

7. Brodtmann, A., Khlif, M. S., Bird, L. J., Cumming, T., & Werden, E. (2021). Hippocampal Volume and Amyloid PET Status Three Years After Ischemic Stroke: A Pilot Study. Journal of Alzheimer’s Disease, 80(2), 527–532. 10.3233/JAD-201525

8. Brodtmann, A., Khlif, M. S., Egorova, N., Veldsman, M., Bird, L. J., & Werden, E. (2020). Dynamic Regional Brain Atrophy Rates in the First Year After Ischemic Stroke. Stroke, 51(9). 10.1161/STROKEAHA.120.030256

9. Brodtmann, A., Pardoe, H., Li, Q., Lichter, R., Ostergaard, L., & Cumming, T. (2012). Changes in regional brain volume three months after stroke. Journal of the Neurological Sciences, 322(1–2), 122–128. 10.1016/j.jns.2012.07.019

10. Brodtmann, A., Werden, E., Khlif, M. S., Bird, L. J., Egorova, N., Veldsman, M., Pardoe, H., Jackson, G., Bradshaw, J., Darby, D., Cumming, T., Churilov, L., & Donnan, G. (2021). Neurodegeneration Over 3 Years Following Ischaemic Stroke: Findings From the Cognition and Neocortical Volume After Stroke Study. Frontiers in Neurology, 12, 754204. 10.3389/fneur.2021.754204

11. Brodtmann, A., Werden, E., Pardoe, H., Li, Q., Jackson, G., Donnan, G., Cowie, T., Bradshaw, J., Darby, D., & Cumming, T. (2014). Charting Cognitive and Volumetric Trajectories after Stroke: Protocol for the Cognition and Neocortical Volume after Stroke (CANVAS) Study. International Journal of Stroke, 9(6), 824–828. 10.1111/ijs.12301

12. Campbell, B. C. V., De Silva, D. A., Macleod, M. R., Coutts, S. B., Schwamm, L. H., Davis, S. M., & Donnan, G. A. (2019). Ischaemic stroke. Nature Reviews Disease Primers, 5(1), 70. 10.1038/s41572-019-0118-8

13. Desikan, R. S., Ségonne, F., Fischl, B., Quinn, B. T., Dickerson, B. C., Blacker, D., Buckner, R. L., Dale, A. M., Maguire, R. P., Hyman, B. T., Albert, M. S., & Killiany, R. J. (2006). An automated labeling system for subdividing the human cerebral cortex on MRI scans into gyral based regions of interest. NeuroImage, 31(3), 968–980. 10.1016/j.neuroimage.2006.01.021

14. Doubal, F. N., MacLullich, A. M. J., Ferguson, K. J., Dennis, M. S., & Wardlaw, J. M. (2010). Enlarged Perivascular Spaces on MRI Are a Feature of Cerebral Small Vessel Disease. Stroke, 41(3), 450–454. 10.1161/STROKEAHA.109.564914

15. Duering, M., Biessels, G. J., Brodtmann, A., Chen, C., Cordonnier, C., De Leeuw, F.-E., Debette, S., Frayne, R., Jouvent, E., Rost, N. S., Ter Telgte, A., Al-Shahi Salman, R., Backes, W. H., Bae, H.-J., Brown, R., Chabriat, H., De Luca, A., deCarli, C., Dewenter, A., … Wardlaw, J. M. (2023). Neuroimaging standards for research into small vessel disease—Advances since 2013. The Lancet Neurology, 22(7), 602–618. 10.1016/S1474-4422(23)00131-X

16. Duperron, M. G., Tzourio, C., Sargurupremraj, M., Mazoyer, B., Soumaré, A., Schilling, S., Amouyel, P., Chauhan, G., Zhu, Y. C., & Debette, S. (2018). Burden of dilated perivascular spaces, an emerging marker of cerebral small vessel disease, is highly heritable. Stroke, 49(2), 282–287. 10.1161/STROKEAHA.117.019309

17. Egorova-Brumley, N., Dhollander, T., Khan, W., Khlif, M. S., Ebaid, D., & Brodtmann, A. (2023). Changes in White Matter Microstructure Over 3 Years in People With and Without Stroke. Neurology, 100(16). 10.1212/WNL.0000000000207065

18. Fonov, V., Evans, A., McKinstry, R., Almli, C., & Collins, D. (2009). Unbiased nonlinear average age-appropriate brain templates from birth to adulthood. NeuroImage, 47, S102. 10.1016/s1053-8119(09)70884-5

19. Francis, F., Ballerini, L., & Wardlaw, J. M. (2019). Perivascular spaces and their associations with risk factors, clinical disorders and neuroimaging features: A systematic review and meta-analysis. International Journal of Stroke, 14(4), 359–371. 10.1177/1747493019830321

20. Goscinski, W. J., McIntosh, P., Felzmann, U., Maksimenko, A., Hall, C. J., Gureyev, T., Thompson, D., Janke, A., Galloway, G., Killeen, N. E. B., Raniga, P., Kaluza, O., Ng, A., Poudel, G., Barnes, D. G., Nguyen, T., Bonnington, P., & Egan, G. F. (2014). The multi-modal Australian ScienceS imaging and visualization environment (MASSIVE) high performance computing infrastructure: Applications in neuroscience and neuroinformatics research. Frontiers in Neuroinformatics, 8(MAR), 30. 10.3389/FNINF.2014.00030/BIBTEX

21. Gu, J., Chen, Y., Tang, H., Chen, X., & Xing, S. (2025). Impaired glymphatic system is associated with secondary neuronal injury in the thalamus following cerebral cortical infarction. Brain Research Bulletin, 224, 111330. 10.1016/j.brainresbull.2025.111330

22. Henschel, L., Conjeti, S., Estrada, S., Diers, K., Fischl, B., & Reuter, M. (2020). FastSurfer—A fast and accurate deep learning based neuroimaging pipeline. NeuroImage, 219. 10.1016/j.neuroimage.2020.117012

23. Hosoki, S., Hansra, G. K., Jayasena, T., Poljak, A., Mather, K. A., Catts, V. S., Rust, R., Sagare, A., Kovacic, J. C., Brodtmann, A., Wallin, A., Zlokovic, B. V., Ihara, M., & Sachdev, P. S. (2023). Molecular biomarkers for vascular cognitive impairment and dementia. Nature Reviews Neurology, 19(12), 737–753. 10.1038/s41582-023-00884-1

24. Iliff, J. J., Wang, M., Liao, Y., Plogg, B. A., Peng, W., Gundersen, G. A., Benveniste, H., Vates, G. E., Deane, R., Goldman, S. A., Nagelhus, E. A., & Nedergaard, M. (2012). A Paravascular Pathway Facilitates CSF Flow Through the Brain Parenchyma and the Clearance of Interstitial Solutes, Including Amyloid β. Science Translational Medicine, 4(147). 10.1126/scitranslmed.3003748

25. Isensee, F., Jaeger, P. F., Kohl, S. A. A., Petersen, J., & Maier-Hein, K. H. (2021). nnU-Net: A self-configuring method for deep learning-based biomedical image segmentation. Nature Methods, 18(2), 203–211. 10.1038/s41592-020-01008-z

26. Johnson, A. C. (2023). Hippocampal Vascular Supply and Its Role in Vascular Cognitive Impairment. Stroke, 54(3), 673–685. 10.1161/STROKEAHA.122.038263

27. Khlif, M. S., Werden, E., Bird, L. J., Egorova[Brumley, N., & Brodtmann, A. (2022). Atrophy of Ipsilesional Hippocampal Subfields Vary Over First Year After Ischemic Stroke. Journal of Magnetic Resonance Imaging, 56(1), 273–281. 10.1002/jmri.28009

28. Lau, K. K., Li, L., Lovelock, C. E., Zamboni, G., Chan, T. T., Chiang, M. F., Lo, K. T., Küker, W., Mak, H. K. F., & Rothwell, P. M. (2017). Clinical Correlates, Ethnic Differences, and Prognostic Implications of Perivascular Spaces in Transient Ischemic Attack and Ischemic Stroke. Stroke, 48(6), 1470–1477. 10.1161/STROKEAHA.117.016694

29. Lau, K. K., Li, L., Schulz, U., Simoni, M., Chan, K. H., Ho, S. L., Cheung, R. T. F., Küker, W., Mak, H. K. F., & Rothwell, P. M. (2017). Total small vessel disease score and risk of recurrent stroke. Neurology, 88(24), 2260–2267. 10.1212/WNL.0000000000004042

30. Lei, H., Wu, X., Ambler, G., Werring, D., Fang, S., Lin, H., Huang, H., Liu, N., & Du, H. (2024). Association between Perivascular Spaces Burden and Future Stroke Risk in Ischemic Stroke and Transient Ischemic Attack: A Systematic Review and Meta-Analysis. European Neurology, 87(3), 130–139. 10.1159/000539730

31. Li, Y., Zhu, Z., Chen, J., Zhang, M., Yang, Y., & Huang, P. (2020). Dilated Perivascular Space in the Midbrain May Reflect Dopamine Neuronal Degeneration in Parkinson’s Disease. Frontiers in Aging Neuroscience, 12(June), 1–7. 10.3389/fnagi.2020.00161

32. Liu, C. F., Hsu, J., Xu, X., Kim, G., Sheppard, S. M., Meier, E. L., Miller, M. I., Hillis, A. E., & Faria, A. V. (2023). Digital 3D Brain MRI Arterial Territories Atlas. Scientific Data, 10(1), 1–17. 10.1038/s41597-022-01923-0

33. Lynch, K. M., Sepehrband, F., Toga, A. W., & Choupan, J. (2023). Brain perivascular space imaging across the human lifespan. NeuroImage, 271, 120009. 10.1016/J.NEUROIMAGE.2023.120009

34. Mestre, H., Du, T., Sweeney, A. M., Liu, G., Samson, A. J., Peng, W., Mortensen, K. N., Stæger, F. F., Bork, P. A. R., Bashford, L., Toro, E. R., Tithof, J., Kelley, D. H., Thomas, J. H., Hjorth, P. G., Martens, E. A., Mehta, R. I., Solis, O., Blinder, P., … Nedergaard, M. (2020). Cerebrospinal fluid influx drives acute ischemic tissue swelling. Science, 367(6483). 10.1126/science.aaw7462

35. Park, C., Shin, N.-Y., Nam, Y., Yoon, U., Ahn, K., & Lee, S.-K. (2023). Characteristics of perivascular space dilatation in normal aging. Human Brain Mapping. 10.1002/HBM.26277

36. Pham, W., Jarema, A., Rim, D., Chen, Z., Khlif, M. S. H., Macefield, V. G., Henderson, L. A., & Brodtmann, A. (2025). *A Comprehensive Framework for Automated Segmentation of Perivascular Spaces in Brain MRI with the nnU-Net* (arXiv:2411.19564). arXiv. 10.48550/arXiv.2411.19564

37. Pham, W., Lynch, M., Spitz, G., O’Brien, T., Vivash, L., Sinclair, B., & Law, M. (2022). A critical guide to the automated quantification of perivascular spaces in magnetic resonance imaging. Frontiers in Neuroscience, 16(December), 1–27. 10.3389/fnins.2022.1021311

38. Potter, G. M., Doubal, F. N., Jackson, C. A., Chappell, F. M., Sudlow, C. L., Dennis, M. S., & Wardlaw, J. M. (2015). Enlarged perivascular spaces and cerebral small vessel disease. International Journal of Stroke, 10(3), 376–381. 10.1111/ijs.12054

39. Ramaswamy, S., Khasiyev, F., & Gutierrez, J. (2022). Brain Enlarged Perivascular Spaces as Imaging Biomarkers of Cerebrovascular Disease: A Clinical Narrative Review. Journal of the American Heart Association, 11(24), e026601. 10.1161/JAHA.122.026601

40. Song, Q., Cheng, Y., Wang, Y., Liu, J., Wei, C., & Liu, M. (2021). Enlarged perivascular spaces and hemorrhagic transformation after acute ischemic stroke. Annals of Translational Medicine, 9(14), 1126–1126. 10.21037/atm-21-1276

41. Tian, Y., Wang, M., Pan, Y., Meng, X., Zhao, X., Liu, L., Wang, Y., & Wang, Y. (2024). In patients who had a stroke or TIA, enlarged perivascular spaces in basal ganglia may cause future haemorrhagic strokes. Stroke and Vascular Neurology, 9(1), 8–17. 10.1136/svn-2022-002157

42. Walt, S. van der, Schönberger, J. L., Nunez-Iglesias, J., Boulogne, F., Warner, J. D., Yager, N., Gouillart, E., & Yu, T. (2014). scikit-image: Image processing in Python. PeerJ, 2, e453. 10.7717/peerj.453

43. Wardlaw, J. M., Smith, E. E., Biessels, G. J., Cordonnier, C., Fazekas, F., Frayne, R., Lindley, R. I., O’brien, J. T., Barkhof, F., Benavente, O. R., Black, S. E., Brayne, C., Breteler, M., Chabriat, H., Decarli, C., De Leeuw, F.-E., Doubal, F., Duering, M., Fox, N. C., … Dichgans, M. (2013). Neuroimaging standards for research into small vessel disease and its contribution to ageing and neurodegeneration. Lancet Neurol, 12(8), 822–838. 10.1016/S1474-4422(13)70124-8

44. Waymont, J. M. J., Valdés Hernández, M. D. C., Bernal, J., Duarte Coello, R., Brown, R., Chappell, F. M., Ballerini, L., & Wardlaw, J. M. (2024). Systematic review and meta-analysis of automated methods for quantifying enlarged perivascular spaces in the brain. NeuroImage, 297, 120685. 10.1016/j.neuroimage.2024.120685

